# Rare biallelic loss-of-function variants in the *LRRK2* kinase cause interstitial lung disease

**DOI:** 10.64898/2026.05.15.26352763

**Authors:** Tugba Kalayci, Kathryn A. Wikenheiser-Brokamp, Sara Gomes, Eshaan Rawat, Anna Peljto, Anthea Cheung, Sevinc Citak, Mustafa Vayvada, M. Angeles Montero-Fernandez, Nesrin Mogulkoc, N. Gulfer Okumus, Ilknur Suer, Joseph Kitzmiller, Cheng-Lun Na, Xinyuan Li, Sheila Bell, Kathleen C. S Cook, Monther Abu-Remaileh, David A. Schwartz, Zehra Oya Uyguner, Jeffrey A Whitsett, Dario R. Alessi, Alexander Zimprich, William J Zacharias, Esther Sammler

## Abstract

We report that biallelic *LRRK2* loss-of-function (LoF) causes a Mendelian form of interstitial lung disease characterized by alveolar epithelial cell dysfunction and lung fibrosis in two brothers with a homozygous nonsense variant. Integrated clinical, imaging, histopathological, and biomarker analyses showed absent LRRK2 protein, reduced Rab10 phosphorylation, impaired alveolar type 2 cell function, and disrupted surfactant homeostasis. Consistent with a recessive genetic disorder, heterozygous LoF carriers have not been reported to have a lung phenotype. Additional biallelic *LRRK2* LoF cases were identified in ILD cohorts, linking LRRK2 LoF to lung disease, in contrast to heterozygous activating missense variants that cause Parkinson’s disease (PD).

## Introduction

Leucine-rich repeat kinase 2 (LRRK2) is a multifunctional protein kinase that regulates intracellular vesicular trafficking through phosphorylation of Rab GTPases and related components of the endolysosomal and secretory pathways. ^1^ Pathogenic gain-of-function (GoF) *LRRK2* variants are amongst the most common genetic causes of Parkinson’s disease (PD) and may contribute to idiopathic PD, making LRRK2 a target for disease-modifying therapies. ^2, 3, 4, 5^ Accordingly, multiple therapeutic strategies to reduce LRRK2 activity including small-molecule kinase inhibitors, antisense oligonucleotides, and targeted protein degraders are in clinical development (https://zenodo.org/records/15604012).

A key translational consideration is the potential for on-target effects that may impact lung tolerability. Human loss-of-function genetics offers a framework to evaluate such risks. Heterozygous *LRRK2* LoF variants, present in approximately 1 in 500 individuals, are not associated with overt pulmonary disease, suggesting partial reduction of LRRK2 activity is tolerated. ^6^ The consequences of biallelic LoF in humans have not been defined. Preclinical *LRRK2* knockout and pharmacological inhibition studies in rodents and non-human primates causes pulmonary alveolar type 2 (AT2) cell abnormalities, impaired lamellar body (LB) homeostasis and histopathological changes suggestive of early interstitial remodeling, raising the possibility that reduced LRRK2 activity disrupts surfactant processing and alveolar epithelial cell function thereby predisposing to interstitial lung disease (ILD). ^7, 8, 9^ Here, we show that biallelic *LRRK2* LoF causes a Mendelian form of ILD characterized by alveolar epithelial dysfunction and lung fibrosis. These findings support a dose-dependent role for LRRK2 in pulmonary homeostasis and inform the therapeutic window and tolerability risks of systemically administered LRRK2-targeted strategies, including those for PD.

## Results

### Familial biallelic *LRRK2* LoF is associated with development of ILD

The index patient (III-2, Fig 1A) is a male individualwho presented with fatigue, progressive dyspnea, exercise intolerance, chronic dry cough, significant weight loss, and digital clubbing with longstanding symptoms. Respiratory symptoms progressed to dyspnea at rest over time, and advanced lung disease ultimately required lung transplantation. Pulmonary function testing (PFT) showed a restrictive pattern (Table S1). High-resolution computed tomography (HRCT) of the chest revealed bilateral emphysematous changes, peripheral bronchiolectasis, septal thickening, and nodular ground-glass opacities consistent with ILD (Fig 1B, *a-c*). Analysis of the explanted lungs revealed characteristic features of usual interstitial pneumonia including temporal and spatial heterogeneity exemplified by ongoing fibrosis in the form of fibroblastic foci at the interface between architecturally distorted end-stage honeycomb lung and relatively preserved lung parenchyma (Fig 1C-E). Enlarged AT2 cells with vacuolated cytoplasm lined thickened alveolar septa in the more preserved lung parenchyma (Fig 1F-G). Electron microscopy showed extensive abnormal LB-like organelles in enlarged AT2 cells (Fig S1). An affected sibling (III-1) developed dyspnea in early adulthood with progression to dyspnea at rest and digital clubbing later in adulthood. HRCT performed during adulthood showed multifocal ground-glass opacities with “crazy paving” predominantly in the lower lobes (Fig 1B, *d-f*). A lung biopsy showed patchy but prominent alveolar septal expansion by loose connective tissue proliferations resembling fibroblastic foci (Fig 1H-J), and epithelial cell hyperplasia consisting of enlarged vacuolated AT2 cells similar to those seen in the index patient’s explant combined with squamous metaplasia indicative of alveolar epithelial injury. Bronchiolectasis with luminal mucostasis was also present (Fig 1H-L). Advanced lung disease was present at latest follow-up. No clear environmental or occupational exposures were identified. ^10^ Two additional siblings (III-3 and III-4), one in the late 20ies and one in late adolescent, were asymptomatic, as were both parents (II-1, and II-2), who were over 60 years of age. Pulmonary evaluation of the parents and the heterozygous sibling, including spirometry and diffusion capacity for carbon monoxide, was normal, and chest radiographs were unremarkable.

**Figure 1.**
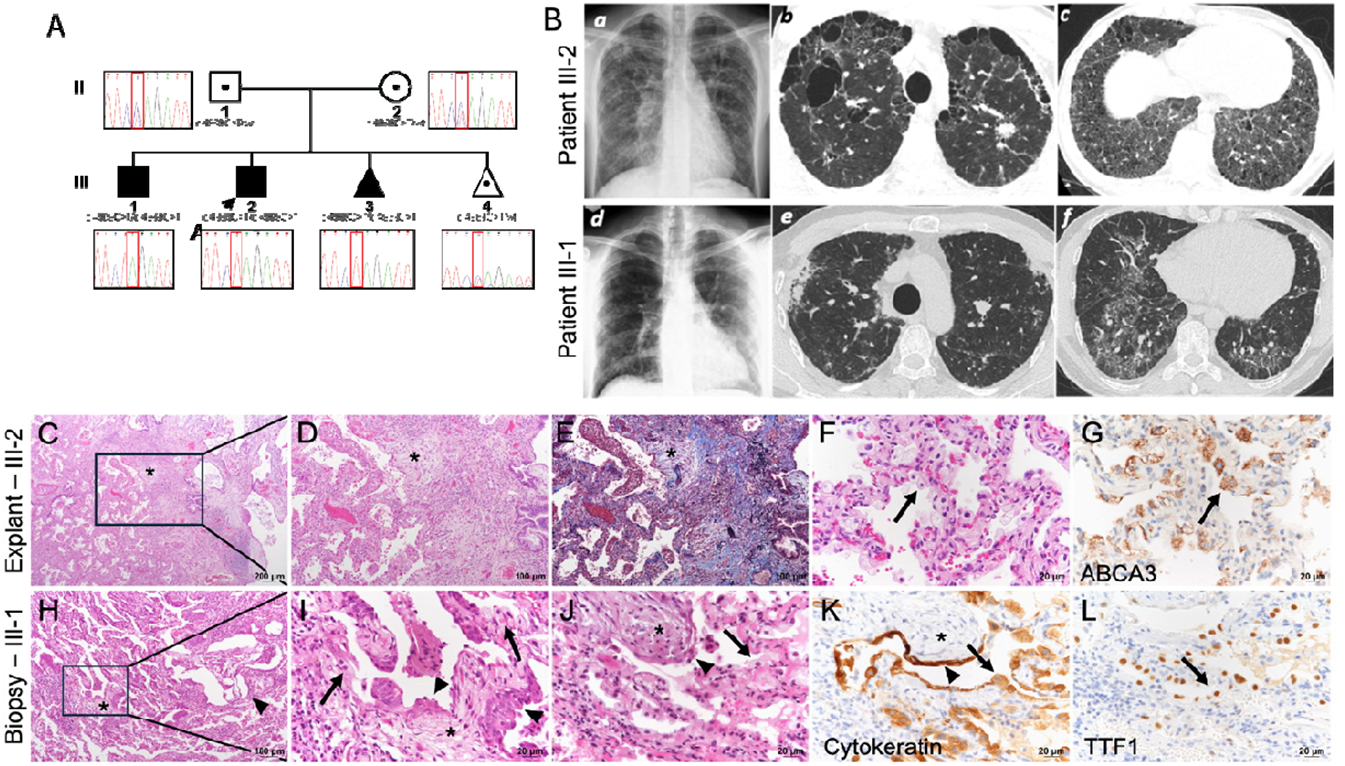
Lung phenotype of a family affected by early-onset ILD associated with biallelic *LRRK2* LoF variants. **A) Pedigree:** Squares represent males; circles represent females. Filled symbols indicate individuals homozygous for the *LRRK2* variant (c.4969C>T, p.Gln1657*). The index patient (III-2) is marked with an arrow. Heterozygous individuals (II-1, II-2, and III-4) are denoted by a dot in the center of the symbol. Genotype information was confirmed by Sanger sequencing and corresponding electropherogram traces are shown under each sequenced individual. A recessive genetic etiology was suspected based on familial clustering. **B. Chest imaging:** Chest radiographs and high-resolution computed tomography (HRCT) images of the proband (III-2) (*a-c*) and affected brother (III-1) (*d-f*). Chest radiographs (a, and d) reveal bilateral reticulonodular opacities. HRCT images show bilateral emphysematous changes, peripheral bronchiolectasis, interlobular septal thickening, and nodular ground-glass opacities in both lungs-findings characteristic of chronic fibrosing ILD. **C-L. Lung Histopathology**: Histologic sections of the explant lungs from the index patient III-2 (C-G) show extensive end stage honeycomb change (C, top right) adjacent to relatively preserved parenchyma with expanded alveolar septa (C, bottom left). Fibroblastic foci (C-E, *) are present at the interface been honeycomb and more preserved lung that are highlighted by Trichrome/VVG stain (E, *). Enlarged alveolar type 2 (AT2) cells with vacuolated cytoplasm line more preserved alveolar septa (F, arrow) and are confirmed as AT2 cells by positive immunostaining for the AT2 cell marker, ABCA3 (G, arrow). The lung biopsy from the affected brother III-1 (H-L) shows bronchiolectasis with mucostasis (H, arrowhead) and alveolar septal expansion by loose connective tissue fibrosis (H-K, *) and epithelial cell hyperplasia consisting of enlarged AT2 cells with vacuolated cytoplasm (I-L, arrows) and squamous metaplasia (I, arrowheads). The flattened injured alveolar epithelial cells overlying the septal fibrosis are highlighted by immunostaining for the epithelial cell marker, pan-cytokeratin (K, arrowhead). The enlarged vacuolated AT2 cells are diffusely immunopositive for both pan-cytokeratin (K, arrow) and the lung epithelial cell marker, TTF1 (L, arrow).

### Genetic analysis reveals homozygous LRRK2 nonsense variant

Given consanguinity, early disease onset, and two affected siblings, a genetic etiology was suspected. Exome sequencing (ES) of the affected brothers did not identify pathogenic variants in known ILD genes (e.g., *SFTPC, SFTPB, ABCA3, MUC5B, BLOC* genes and *NKX2-1*). ^10, 11^ Both affected individuals carried a homozygous nonsense variant in *LRRK2* (NM 198578.3: c.4969C>T; p.Gln1657*), introducing a premature stop codon within the COR-B domain truncating the protein upstream of the kinase domain. The same homozygous variant was present in the asymptomatic sibling in the late 20ies who declined clinical evaluation. Both parents and the sister were heterozygous carriers (Fig 1A). This variant is extremely rare in gnomAD (v4.1), observed twice among 1,611,666 alleles and exclusively in the heterozygous state. ^12^

### LRRK2 p. Gln1657* disrupts LRRK2 protein expression and kinase activity *in vivo*

LRRK2 kinase pathway activity was assessed in peripheral blood neutrophils from the two affected brothers, their siblings, and parents. LRRK2 protein was undetectable in homozygous LoF individuals (Fig 2A). Phosphorylation of the LRRK2 substrate Rab10 at Thr73 (pThr73-Rab10) and LRRK2 phosphorylation at Ser910 and Ser935 were absent (Fig 2B-D). Heterozygous family members exhibited detectable LRRK2 protein and preserved phosphorylation (Fig 2A-D). *Ex vivo* treatment of neutrophils with the selective LRRK2 inhibitor MLi-2 abolished phosphorylation of Rab10 and LRRK2, confirming functional *LRRK2* kinase activity in heterozygotes (Fig 2B-D). Unrelated healthy controls are shown for comparison (Fig 2A-D). Urinary bis(monoacylglycero)phosphate (BMP), a biomarker of LRRK2 kinase activation and lysosomal dysfunction, ^13^ was reduced in homozygous patients compared to heterozygous individuals and controls, supporting *in vivo* pathway inactivation (Fig 2E).

**Figure 2:**
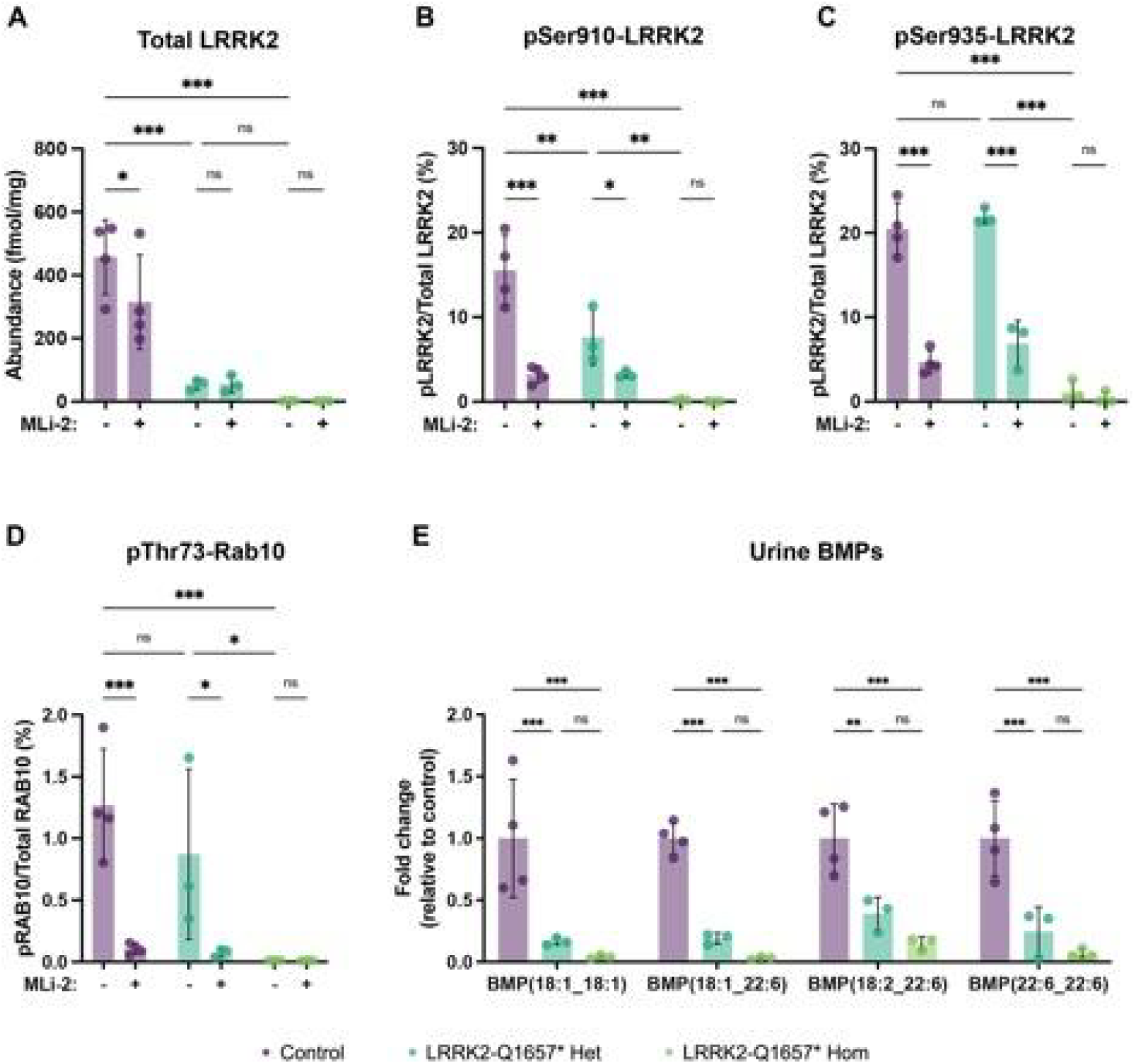
LRRK2 biomarker profile of carriers of the LRRK2 LoF p.Gln1657* variant. **(A-D)** LRRK2 and pThr73-Rab10 peptide abundance is significantly reduced in neutrophils of heterozygous and homozygous carriers of the LRRK2 p.Gln1657* variant. Levels of total LRRK2, pSer910-LRRK2, pSer935LRRK2, pThr73-Rab10 and total Rab10 were measured by targeted mass spectrometry in protein extracts from neutrophils treated with DMSO (-) or specific LRRK2 kinase inhibitor 200 nM MLi-2 (+). **(A)** Total LRRK2 levels are shown as abundance per milligram of protein extract. **(B)** pSer910-LRRK2 and **(C)** pSer935-LRRK2 levels are expressed as a percentage of total LRRK2. **(D)** pThr73-Rab levels are expressed as a percentage of total Rab10. **(A-D)** Data presented as mean ± SD. **(E)** BMPs are significantly reduced in the urine of heterozygous and homozygous carriers of the LRRK2 p.Gln1657* variant. Targeted lipidomics data presented as mean ± SEM, with fold change relative to mean of control samples. BMP levels endogenously normalized by PC (18:1_18:1). **(A-E)** All analyses were performed on unrelated healthy controls (n = 4), heterozygous (n = 3) and homozygous (n = 3) carriers of the LRRK2 p.Gln1657* variant. Statistical analysis was performed using ordinary two-way ANOVA with post-hoc Tukey’s multiple comparisons test (*p<0.05, **p<0.01, ***p<0.001).

### LRRK2 deficiency is associated with aberrant lipid processing in AT2 cells

We examined explanted lung tissue from the index patient and biopsy tissue from his affected brother to determine the cellular biology underlying the ILD phenotype. AT2 cells synthesize, secrete, and recycle surfactant proteins and lipids and express high levels of LRRK2. ^9, 14,15^ Additionally, many genetic causes of ILD converge on AT2 cell dysfunction. ^16, 17, 18^ To define AT2 cell state in LRRK2 LoF, we evaluated pro-surfactant protein C (pSFTPC), mature surfactant protein B (mSFTB) and LB markers, ABCA3 and LAMP3, by super resolution microscopy (Fig 3A-F). In normal AT2 cells, ABCA3 and LAMP3 stain LB membranes, pSFTPC localizes perinuclearly, and mSFTPC is found in a heterogeneous pattern within LB. AT2 cells in lungs of *LRRK2* LoF individuals showed marked morphological abnormalities, with mislocalized pSFTPC, prominent intracellular LB-like organelles with heterogeneous LAMP3 and ABCA3 staining, loss of organized LB structure, and mislocalization of mSFTPC outside of LB-like organelles. This enlarged, vacuolated AT2 cell phenotype recapitulated findings in LRRK2 knock-out animals and animals treated with LRRK2 inhibitors. ^7, 8, 9^ The accumulation of pro-fibrotic epithelial cells marked by loss of SFTPC and high-level expression of the cytokeratins KRT8 and KRT17 is a hallmark of multiple genetic and sporadic human ILDs. ^19,20,21,22^ Concordantly, abundant KRT8^high^/KRT17^low^, KRT8^high^/KRT17^high^, and KRT8^low^/KRT17^high^ epithelial cells were found throughout more severely fibrotic regions of both LRRK2 LoF patient lungs (Fig 3G-I). These data indicate that loss of LRRK2 disrupts AT2 cell homeostasis and contributes to profibrotic transition leading to ILD.

**Figure 3:**
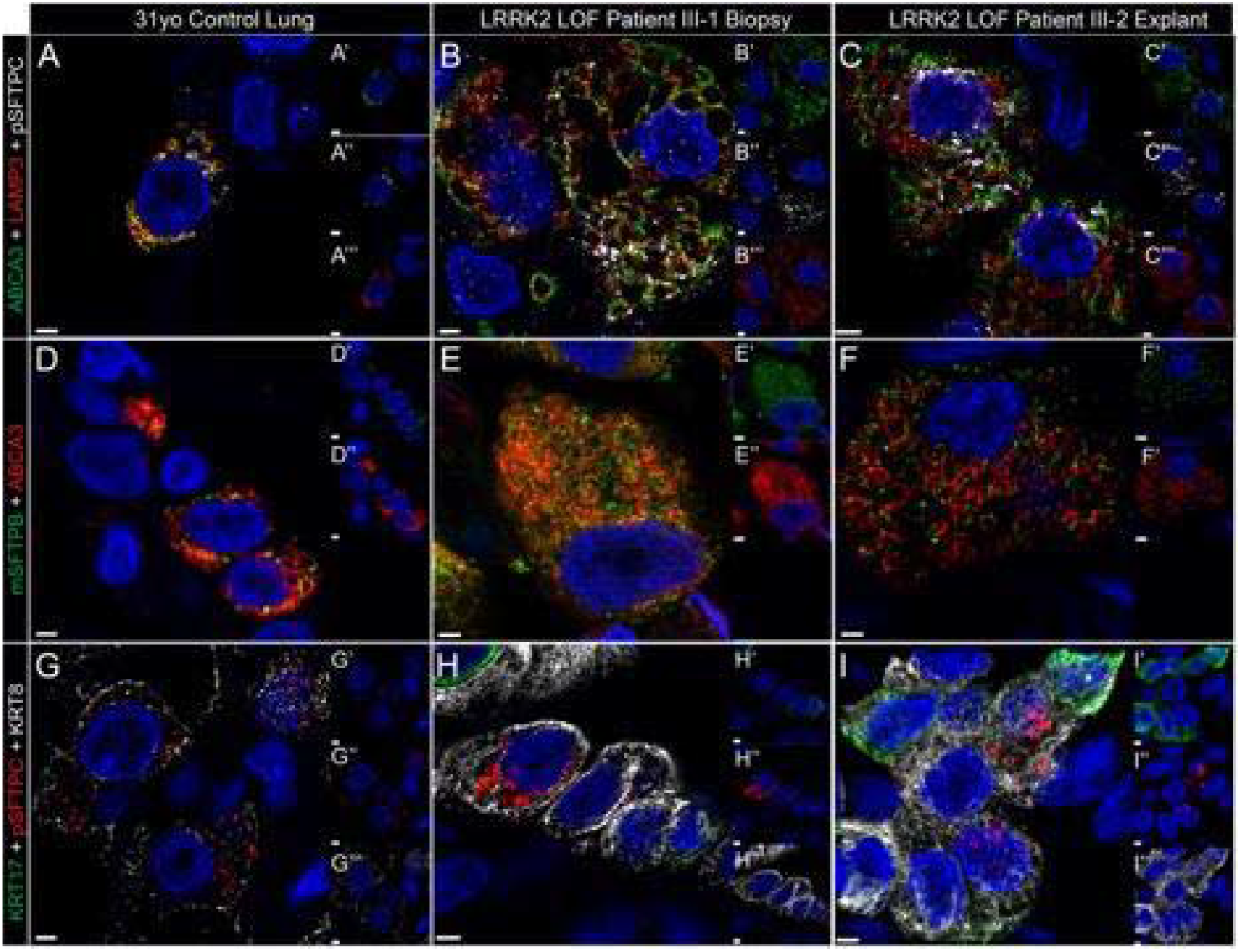
LRRK2 LoF variants drive fibrotic lung remodeling associated with AT2 cell dysfunction, failure of surfactant processing, and cell state transition to basaloid epithelial cell state. Examination of LRRK2 LoF patients and age-matched control lung samples for the lamellar body proteins ABCA3 (A-F) and LAMP3 (A-C), the surfactant proteins pro-SFTPC (A-C) and mature SFTPB (D-F), and the fibrosis-associated basaloid markers KRT8 and KRT17 (G-I). Scale bars = 2 microns. Insets show single channel views of each stain.

### Genetic identification of unrelated patients with LRRK2 LoF and ILD

We queried and examined large-scale public genetic databases to identify other potential cases of ILD associated with LRRK2 mutations. In TOPMed (whole-genome sequencing (WGS) of 3,682 idiopathic pulmonary fibrosis (IPF) patients and 2,449 controls), ^23^ we identified a female in her 60ies with ILD carrying a homozygous nonsense *LRRK2* variant (c.6187_6191del; p.Leu2063*) (Tab S1 and Supplementary Material). She was a 1.75 pack-year smoker with dyspnea, PFTs demonstrating a restrictive pattern, and CT imaging showing fibrotic changes with moderate honeycombing. The Leu2063* variant appears 451 times in gnomAD, ^12^ but only in the heterozygous state. This variant is predicted to truncate the kinase domain and abolish activity. In the NIH All of Us Research Program (short-read WGS from 245,388 individuals), ^24^ one ILD patient among 4,299 cases was identified as carrying two distinct LRRK2 LoF variants: a canonical splice donor site mutation at the exon 23/intron 24 junction (c.3096+1G>A), and a nonsense variant (*c*.*3939T>A*; p.Cys1313*), predicted to truncate the LRR domain and eliminate the downstream kinase domain (Tab S1). It is unclear if these variants were in cis or in trans.

### *LRRK2* LoF variants in unrelated patients result in loss of LRRK2 kinase activity

To assess the functional consequences of the identified human *LRRK2* variants, we performed overexpression studies in HEK293 cells. Expression of p.Gln1657* failed to induce phosphorylation of LRRK2 substrate (Fig S2). Similarly, the variants p.Gln1657*, p.Leu2063*, and p.Cys1313* produced truncated proteins with reduced molecular weight and were unable to phosphorylate Rab10 (Fig S2). Collectively, these findings indicate that all the identified *LRRK2* variants identified in unrelated patients result in loss of LRRK2 function.

## Discussion

Here we provide human genetic evidence implicating biallelic *LRRK2* LoF variants as a cause of familial fibrotic ILD in two affected siblings, an unrelated individual with a distinct homozygous *LRRK2* LoF variant, and a fourth individual with possible compound heterozygous LoF variants. Functional characterization in HEK293 cells demonstrated loss of kinase activity across all nonsense variants. In clinical samples from affected family members, homozygous p.Gln1657* carriers lacked detectable LRRK2 protein, exhibited reduced Rab10 phosphorylation, and had decreased urinary BMP levels, indicating systemic LRRK2 deficiency.

Mechanistically, our data support a model in which impaired vesicular trafficking and endolysosomal exocytosis in AT2 cells drive ILD pathogenesis. Disruption of surfactant processing and resulting aberrant lamellar body generation likely leads to epithelial injury, alveolar damage, and activation of pro-fibrotic pathways. This is consistent with existing genetic and biological evidence implicating a causal role for AT2 dysfunction in early-onset ILD, including variants affecting surfactant homeostasis (e.g. SFTPC, ABCA3), lysosome-related trafficking (LAMP3), and disorders of endolysosomal function such as Hermansky-Pudlak syndrome. ^11, 25^ The emergence of pro-fibrotic epithelial cell states in our patients further supports a model of epithelial stress and maladaptive repair contributing to fibrosis.

Together, these data expand the phenotypic spectrum of LRRK2-associated disease beyond PD and establish LRRK2 as a gene where both gain and loss-of-function cause distinct human pathologies. These findings provide new insight into the role of LRRK2 in lung epithelial cell and lysosomal biology, highlight the importance of defining the therapeutic window and tolerability of LRRK2 inhibition or degradation in PD and suggest that activation of the LRRK2 pathway may warrant exploration in fibrotic lung disease. More broadly, this study illustrates how human genetics can inform both the safety and therapeutic targeting of biologically tractable pathways across organ systems.

## Data Availability

All data produced in the present study are available upon reasonable request to the authors. We are in the process of making all data available via Zenodo.

## Acknowledgements

We are deeply grateful to all patients, families, and healthy participants who contributed to this study. We thank our funders, collaborators, and colleagues for valuable discussions, critical feedback, and manuscript review.

This work was supported by the MRC Protein Phosphorylation and Ubiquitylation Unit (MRC PPU), University of Dundee, UK, including the MRC PPU Reagents and Services teams and the MRC PPU Mass Spectrometry Facility for technical and proteomic support.

Donor control lung tissue was obtained from the LifeCenter Organ Donor Network (Cincinnati, OH). We also acknowledge support from the Bio-Imaging and Analysis Facility and the Integrated Pathology Research Facility at Cincinnati Children’s Research Foundation (Cincinnati, OH).

## References

1. Alessi DR, Pfeffer SR. Leucine-Rich Repeat Kinases. Annu Rev Biochem 2024;93(1):261–287. DOI: 10.1146/annurev-biochem-030122-051144.

2. Kruger C, Lim SY, Buhrmann A, et al. Updated MDSGene review on the clinical and genetic spectrum of LRRK2 variants in Parkinson s disease. NPJ Parkinsons Dis 2025;11(1):30. DOI: 10.1038/s41531-025-00881-9.

3. Pitz V, Makarious MB, Bandres-Ciga S, et al. Analysis of rare Parkinson’s disease variants in millions of people. NPJ Parkinsons Dis 2024;10(1):11. DOI: 10.1038/s41531-023-00608-8.

4. Alessi DR, Sammler E. LRRK2 kinase in Parkinson’s disease. Science 2018;360(6384):36–37. DOI: 10.1126/science.aar5683.

5. Lang AE, Hauser RA, Kalia LV, et al. LRRK2 as a Potential Disease-Modifying Target in Sporadic Parkinson’s Disease. Mov Disord 2026;41(2):297–314. DOI: 10.1002/mds.70100.

6. Whiffin N, Armean IM, Kleinman A, et al. The effect of LRRK2 loss-of-function variants in humans. Nat Med 2020;26(6):869–877. DOI: 10.1038/s41591-020-0893-5.

7. Bryce DK, Ware CM, Woodhouse JD, et al. Characterization of the Onset, Progression, and Reversibility of Morphological Changes in Mouse Lung after Pharmacological Inhibition of Leucine-Rich Kinase 2 Kinase Activity. J Pharmacol Exp Ther 2021;377(1):11–19. DOI: 10.1124/jpet.120.000217.

8. Baptista MA, Dave KD, Frasier MA, et al. Loss of leucine-rich repeat kinase 2 (LRRK2) in rats leads to progressive abnormal phenotypes in peripheral organs. PLoS One 2013;8(11):e80705. DOI: 10.1371/journal.pone.0080705.

9. Fuji RN, Flagella M, Baca M, et al. Effect of selective LRRK2 kinase inhibition on nonhuman primate lung. Sci Transl Med 2015;7(273):273ra15. DOI: 10.1126/scitranslmed.aaa3634.

10. Tirelli C, Pesenti C, Miozzo M, Mondoni M, Fontana L, Centanni S. The Genetic and Epigenetic Footprint in Idiopathic Pulmonary Fibrosis and Familial Pulmonary Fibrosis: A State-of-the-Art Review. Diagnostics (Basel) 2022;12(12). DOI: 10.3390/diagnostics12123107.

11. Bridges JP, Vladar EK, Kurche JS, et al. Progressive lung fibrosis: reprogramming a genetically vulnerable bronchoalveolar epithelium. J Clin Invest 2025;135(1). DOI: 10.1172/JCI183836.

12. Chen S, Francioli LC, Goodrich JK, et al. A genomic mutational constraint map using variation in 76,156 human genomes. Nature 2024;625(7993):92–100. DOI: 10.1038/s41586-023-06045-0.

13. Gomes S, Garrido A, Tonelli F, et al. Elevated urine BMP phospholipids in LRRK2 and VPS35 mutation carriers with and without Parkinson’s disease. NPJ Parkinsons Dis 2023;9(1):52. DOI: 10.1038/s41531-023-00482-4.

14. Herzig MC, Kolly C, Persohn E, et al. LRRK2 protein levels are determined by kinase function and are crucial for kidney and lung homeostasis in mice. Hum Mol Genet 2011;20(21):4209–23. DOI: 10.1093/hmg/ddr348.

15. Harney J, Bajaj P, Finley JE, et al. An in vitro alveolar epithelial cell model recapitulates LRRK2 inhibitor-induced increases in lamellar body size observed in preclinical models. Toxicol In Vitro 2021;70:105012. DOI: 10.1016/j.tiv.2020.105012.

16. Whitsett JA, Wert SE, Weaver TE. Diseases of pulmonary surfactant homeostasis. Annu Rev Pathol 2015;10:371–93. DOI: 10.1146/annurev-pathol-012513-104644.

17. Nogee LM. Genetic causes of surfactant protein abnormalities. Curr Opin Pediatr 2019;31(3):330–339. DOI: 10.1097/MOP.0000000000000751.

18. Keehan LA, Ono-Minagi H, Hadhud M, et al. Biallelic LAMP3 variants in 5 families with interstitial lung disease: Evidence of a disease-gene association. Genet Med 2026;28(4):102531. DOI: 10.1016/j.gim.2026.102531.

19. Habermann AC, Gutierrez AJ, Bui LT, et al. Single-cell RNA sequencing reveals profibrotic roles of distinct epithelial and mesenchymal lineages in pulmonary fibrosis. Sci Adv 2020;6(28):eaba1972. DOI: 10.1126/sciadv.aba1972.

20. Wang F, Ting C, Riemondy KA, et al. Regulation of epithelial transitional states in murine and human pulmonary fibrosis. J Clin Invest 2023;133(22). DOI: 10.1172/JCI165612.

21. Ting C, Konopka K, Benedeck RE, et al. Biomarkers Unveil Insights into Pathology of Transitional Epithelial States in Pulmonary Fibrosis. Am J Respir Crit Care Med 2024;210(5):687–690. DOI: 10.1164/rccm.202310-1839LE.

22. DePianto DJ, Heiden JAV, Morshead KB, et al. Molecular mapping of interstitial lung disease reveals a phenotypically distinct senescent basal epithelial cell population. JCI Insight 2021;6(8). DOI: 10.1172/jci.insight.143626.

23. Peljto AL, Blumhagen RZ, Walts AD, et al. Idiopathic Pulmonary Fibrosis Is Associated with Common Genetic Variants and Limited Rare Variants. Am J Respir Crit Care Med 2023;207(9):1194–1202. DOI: 10.1164/rccm.202207-1331OC.

24. All of Us Research Program Genomics I. Genomic data in the All of Us Research Program. Nature 2024;627(8003):340–346. DOI: 10.1038/s41586-023-06957-x.

25. Yokoyama T, Gochuico BR. Hermansky-Pudlak syndrome pulmonary fibrosis: a rare inherited interstitial lung disease. Eur Respir Rev 2021;30(159). DOI: 10.1183/16000617.0193-2020.

