## Supplementary Material for "Rare biallelic loss-of-function variants in the *LRRK2* kinase cause interstitial lung disease"

**Table of Contents**

1. Clinical Findings
 2. Supplementary Figures
 3. Supplementary Tables
 4. Materials and Methods
 5. References

**1. Clinical Findings**

**Detailed Clinical Description of the Index Family**

The index patient (III-2) was an adult male who presented with fatigue, progressive dyspnea, exercise intolerance, chronic dry cough, marked weight loss, and digital clubbing with longstanding symptoms. Physical examination showed digital clubbing and bibasilar Velcro crackles.

Pulmonary function testing demonstrated a restrictive ventilatory defect with preserved airflow: FEV₁ 3440 mL (83% predicted), FVC 3820 mL (76% predicted), and FEV₁/FVC ratio 90.06% (111% predicted). TLC was moderately reduced (5210 mL, 74% predicted). DLCO was severely reduced (6.58 mL/mmHg/min, 19% predicted), and DLCO/VA was 1.30 mL/mmHg/min/L (26% predicted). The elevated FVC%/DLCO% ratio (4.0) indicated disproportionate impairment of gas exchange relative to lung volumes.

During the six-minute walk test, SpO₂ decreased from 95% to 82% (heart rate 64/min to 95/min; Borg 1 to 6), indicating exertional desaturation. At 1 minute of recovery, SpO₂ was 89% and heart rate 78/min (pulse recovery, 17/min). Functional status indicated moderate limitation (score, 50/100). HRCT showed bilateral emphysema, peripheral bronchiolectasis, septal thickening, and nodular ground-glass opacities, consistent with interstitial lung disease.

An affected sibling (III-1), a male individual in his late 30ies, developed progressive dyspnea beginning in early adulthood, progressing to dyspnea at rest and digital clubbing during adulthood. Computed tomography later raised suspicion for interstitial lung disease. Auscultation revealed occasional crackles. Digital clubbing was not observed at that time.

Pulmonary function testing showed a marked restrictive pattern: FEV₁ 2270 mL (50% predicted), FVC 2640 mL (47% predicted), and FEV₁/FVC ratio 86.16% (107% predicted). TLC was severely reduced (3100 mL, 38% predicted). DLCO was 4.08 mL/mmHg/min (32% predicted), whereas DLCO/VA was relatively preserved (1.39 mL/mmHg/min/L, 88% predicted), suggesting reduced alveolar surface area with preserved efficiency per unit volume. The FVC%/DLCO% ratio was 1.47. Serial measurements demonstrated disease progression, with an FVC decline of 340 mL (11.4%) over one year.

During exercise testing, SpO₂ decreased from 97% to 87% (heart rate 89/min to 121/min; Borg 0 to 4), consistent with exertional hypoxemia. HRCT showed multifocal ground-glass opacities with “crazy paving,” suggestive of an active inflammatory process. The lung biopsy was initially interpreted as showing patchy subpleural and centrilobular areas of increased interstitial cellularity, peribronchiolar metaplasia, foci of organizing pneumonia, and alveolar spaces lined by finely vacuolated and enlarged pneumocytes highlighted by cytokeratin AE1/AE3 immunostain. The features were interpreted as suspicious for a lysosomal storage disease and exclusion of a genetic alteration was recommended.

Additional relatives carrying heterozygous and in one case homozygous variants were asymptomatic. Pulmonary evaluation of the heterozygous carriers, including spirometry and DLCO, was normal. Chest radiographs were unremarkable; therefore, HRCT was not performed.

**An additional relative carrying the variant declined further clinical evaluation.** No family member had a history of smoking or relevant environmental or occupational exposure.

**Additional Case from Population Database**

In the TOPMed database, ^1^ one older adult female in her sixties with a limited smoking history had interstitial lung disease diagnosed in her sixth decade.

Pulmonary function testing showed FVC 2380 mL (73% predicted), FEV₁/FVC ratio 91%, and TLC 3080 mL (59% predicted). DLCO was moderately to severely reduced (10.68 mL/mmHg/min, 50% predicted), whereas DLCO/VA was relatively preserved (4.31 mL/mmHg/min/L, 82% predicted). The FVC%/DLCO% ratio was 1.46. Imaging showed moderate honeycombing, consistent with pulmonary fibrosis.

**2. Supplementary Figures**

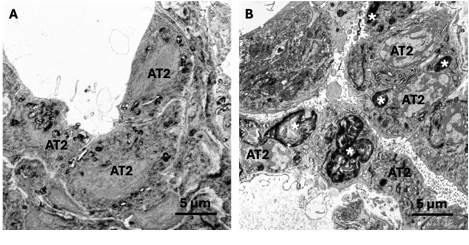

**Figure S1: Ultrastructural abnormalities in alveolar type 2 cells associated with bi-allelic LRRK2 loss-of-function.** Transmission electron microscopy of lung tissue from **(A)** an age matched adult male control donor without lung disease and **(B)** patient III-2 with biallelic *LRRK2* loss-of-function (LoF) variants. In the patient sample, alveolar type 2 (AT2) cells contain aberrant lamellar body (LB)-like organelles (asterisks) that are enlarged, have abnormal fusion, and are associated with lipid accumulation as compared to the smaller, organized LBs in the control donor lung. AT2 cell hyperplasia and increased collagen deposition are also present in the patient lung. The accumulation of aberrant LB-like organelles in the patient lung is consistent with the light microscopic finding of AT2 cell vacuolization (see Fig. 1C–G). Scale bar = 5 μm.

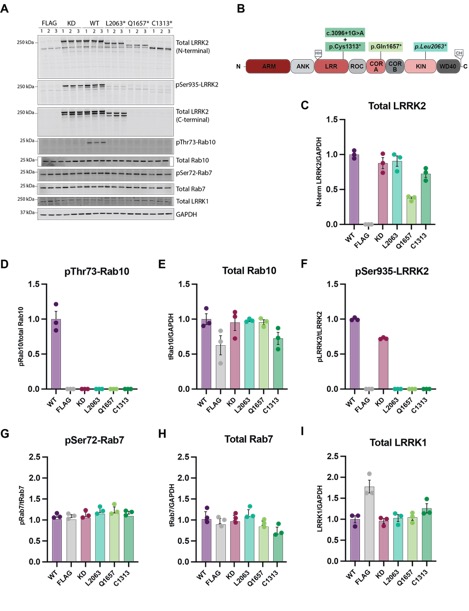

**Figure S2. *LRRK2* LoF variants identified in unrelated patients** **impair kinase activity and downstream Rab10 phosphorylation in HEK293 overexpression assays.** *LRRK2* LoF variants (p.Leu2063*, p.Gln1657*, and p.C1313*) were compared with wild-type (WT) LRRK2 in an established HEK293 overexpression assay. **(A)** Representative quantitative immunoblots from three independent biological replicates with each lane representing an independent biological replicate. LRRK2 kinase activity was assessed by phosphorylation of endogenous Rab10 at threonine 73 (pThr73-Rab10). **(B)** Schematic representation of the variant position within the LRRK2 domain structure. **(C–I)** Quantification of immunoblot signals, expressed as fold change relative to WT. Total LRRK2 was normalized to GAPDH (C), pSer935-LRRK2 normalized to total LRRK2 (D), pThr73-Rab10 normalized to total Rab10 (E), total Rab10 normalized to GAPDH **(F),** total LRRK1 normalized to GAPDH **(G),** pSer72-Rab7 normalized to total Rab7 (H), and total Rab7 normalized to GAPDH (I). Data in (C–I) are presented as means ±SD from three independent biological replicates.

**3. Supplementary Tables**

**Table S1. Clinical and Molecular Characteristics of Patients with Interstitial Lung Disease.**

|  | **Affected relative**  **(III-1)** | **Index patient**  **(III-2)** | **Additional relative**  **(III-3)** | **TOPMed** | | **AllofUs** |
| --- | --- | --- | --- | --- | --- | --- |
| **ILD diagnosis** | **Yes** | **Yes** | **No** | **Yes** | | **Yes** |
| **Symptoms (approximate Period of Onset)** | | | | | | |
| **Fatigue** | **20ies** | **Early onset** | **-** | **NA** | | **NA** |
| **Chronic dry cough** | **-** | **Early onset** | **-** | **Adulthood** | | **NA** |
| **Exercise intolerance** | **20ies** | **Early onset** | **-** | **NA** | | **NA** |
| **Digital clubbing** | **4^th^ decade** | **Early onset** | **-** | **NA** | | **NA** |
| **Dyspnea during rest** | **4^th^ decade** | **Early adulthood** | **-** | **NA** | | **NA** |
| **Pulmonary hypertension** | **-** | **Early adulthood** | **NA** | **NA** | | **NA** |
| **Need of O2 support** | **-** | **Adulthood** | **-** | **NA** | | **NA** |
| **Pulmonary Function** | | | | | | |
| **FEV1** | **2270 ml (50%)** | **3440 ml (83%)** | **NA** | **2150 ml (66%)** | | **NA** |
| **FVC** | **2640 ml (47%)** | **3820 ml (76%)** | **NA** | **2380 ml (73%)** | | **NA** |
| **FEV1/FVC** | **107%** | **111%** | **NA** | **91%** | | **NA** |
| **DLCO** | **32%** | **19%** | **NA** | **50%** | | **NA** |
| **DLCO/VA** | **88%** | **26%** | **NA** | **82%** | | **NA** |
| **FVC%/DLCO%** | **1.47** | **4.0** | **NA** | **1.46** | | **NA** |
| **TLC** | **38%** | **74%** | **NA** | **59%** | | **NA** |
| **Genetic Results for LRRK2** | | | | | | |
| **Domain** | COR-B | COR-B | COR-B | Kinase | | LRR |
| **Variant** | c.4969C>T | c.4969C>T | c.4969C>T | c.6187_6191del | c.3096+1G>A and  c.3939T>A | |
| **Protein** | p.Gln1657* | p.Gln1657* | p.Gln1657* | p.Leu2063* | splice and p.Cys1313* | |
| **Zygosity** | Hom | Hom | Hom | Hom | NA | |
| **Frequency (gnomAD_v4.1)** | 2/1,611,666 | 2/1,611,666 | 2/1,611,666 | 451/1,613,786 | 2/249,556 and  17/ 249,378 | |

**Table S1. Clinical and Molecular Characteristics of Patients with Interstitial Lung Disease.** Data are shown for multiple family members with interstitial lung disease (affected brother [III-1], index patient [III-2], and other brother [III-3]) and for individuals identified in the TOPMed ^1^ and All of Us ^2^ databases.

Symptoms are reported with age at onset (in years) when available. Pulmonary function test results for FEV₁ and FVC are presented as absolute values (milliliters) with percentages of predicted values in parentheses and all other pulmonary function parameters are presented as percentages of predicted values. Genetic data include *LRRK2* (NM_198578.4) variants, with domain location, nucleotide and protein changes, zygosity, and allele frequencies from gnomAD (version 4.1). ^3^

**Abbreviations:** DLCO, diffusing capacity of the lung for carbon monoxide; DLCO/VA, diffusing capacity adjusted for alveolar volume; FEV₁, forced expiratory volume in 1 second; FVC, forced vital capacity; TLC, total lung capacity; NA, not available; Hom, homozygous.

**Table S2. Heavy-Labelled Peptides**

| **Target** | **Sequence** |
| --- | --- |
| pThr73-Rab10 | FHpTITTSYYR* |
| Rab10 | NIDEHANEDVER* |
| pSer910-LRRK2 | SNpSISVGEFYR* |
| pSer935-LRRK2 | HSNpSLGPIFDHEDLLK* |
| LRRK2 | IGDEDGHFPAHR* |

* Denotes heavy-labelled amino acids, either R[^13^C_6_ ^15^N_4_] or K[^13^C_6_ ^15^N_2_].

**Table S3. Antibodies used for immunoprecipitation-based enrichment.**

| **Antibody Target** | **Supplier** | **Cat. number** | **Host species** |
| --- | --- | --- | --- |
| pSer935-LRRK2 | Abcam | ab133450 | Rabbit |
| pSer910-LRRK2 | Abcam | ab203181 | Rabbit |
| Total LRRK2 | Antibodies Inc./NeuroMab | 75-253 | Mouse |
| pan-pThr-Rab | Abcam | 230261 | Rabbit |

### **Table S4. Plasmids used for LRRK2 overexpression in HEK293 cells.**

| **DU Number** | **Construct** | **Plasmid** |
| --- | --- | --- |
| DU41799 | FLAG-Empty | pCMV5 |
| DU10128 | FLAG-LRRK2 D2017A (Kinase inactive) | pCMV5 |
| DU6841 | FLAG-LRRK2 Wild type | pCMV5 |
| DU80365 | FLAG-LRRK2 L2063 Stop | pCMV5 |
| DU76258 | FLAG-LRRK2 Q1657 Stop | pCMV5 |
| DU80360 | FLAG-LRRK2 C1313 Stop | pCMV5 |

**Table S5. Antibodies used for immunoblotting.**

| **Antibody Target** | **Supplier** | **Cat. number** | **Host species** | **Dilution** |
| --- | --- | --- | --- | --- |
| pSer935-LRRK2 | Abcam | ab133450 | Rabbit | 1 µg/mL |
| Total LRRK2 | Antibodies Inc./NeuroMab | 75-253 | Mouse | 1 µg/mL |
| pThr73-Rab10 | Abcam | 230261 | Rabbit | 1 µg/mL |
| Total Rab10 | Nanotools | 0680-100 | Mouse | 1 µg/mL |
| pSer92-Rab7 | Abcam | 302494 | Rabbit | 1 µg/mL |
| Total Rab7 | Sigma-Aldrich | R8779 | Mouse | 1 µg/mL |
| LRRK1 | Abcam | 228666 | Rabbit | 1 µg/mL |
| GAPDH | Santa Cruz | sc-32233 | Mouse | 1:5000 |

**Table S6. Antibodies used for immunofluorescence.**

| **Antibody Target** | **Supplier** | **Cat. number** | **Host species** | **Dilution** |
| --- | --- | --- | --- | --- |
| ABCA3 | Seven Hills Bioreagents | WMAB-ABCA3-17  RRID: AB_577285 | Mouse monoclonal | 1:100 |
| KRT8 | Developmental Studies Hybridoma Bank | TROMA-1  RRID: AB_531826 | Mouse monoclonal | 1:100 |
| KRT17 | Santa Cruz | SC-393002  RRID: AB_2893006 | Mouse monoclonal | 1:100 |
| mature SFTPB | In house, JA Whitsett laboratory | In house, GP20 | Guinea pig polyclonal | 1:100 |
| pro-SFTPC | R&D Systems | WRAB 9337  RRID: AB_2335890 | Rabbit polyclonal | 1:500 |

### **4. Materials and Methods**

**Ethics statement and data access:**

The study was approved by the local institutional review board and conducted in accordance with the Declaration of Helsinki. Written informed consent was obtained from all participants. Use of All of Us data was approved by the All of Us Resource Access Board, with an exception granted under the Data and Statistics Dissemination Policy.

**Genetic analysis:**

**Exome sequencing (ES):** Genomic DNA was extracted from peripheral blood samples using standard protocols. ES was performed using the Sophia Genetics Twist HCExome_v2 Kit (Sophia Genetics, Switzerland) for target enrichment. Library preparation was conducted according to the manufacturer’s protocol, including DNA fragmentation, end repair, A-tailing, adapter ligation, and PCR amplification. The enriched libraries were then sequenced on an Illumina NextSeq 6000 platform, generating 150 bp paired-end reads. The sequencing run aimed to achieve an average coverage of at least 100x to ensure high sensitivity for variant detection. Raw sequencing data were processed using Sophia Genetics' DDM (Data Driven Medicine) platform, including read alignment to the GRCh38/hg38 human reference genome, duplicate marking, and variant calling. Variant annotation and filtering were performed according to established clinical guidelines. ^4^

Targeted analysis of genes associated with interstitial lung disease (e.g. *ABCA3, AP3B1, AP3D1, BLOC1S1, BLOC1S2, BLOC1S3, BLOC1S4, BLOC1S5, BLOC1S6, BTNL2, CSF2RA, CSF2RB, DBNDD2, DKC1, DTNBP1, ELMOD2, FAM111B, FARSA, FARSB, HPS1, HPS3, HPS4, HPS5, HPS6, ITGA3, MCM4, MUC5B, NAF1, NKX2-1, NOP10, PARN, POT1, RNF168, RPA1, RTEL1, SFTPA2, SFTPB, SFTPC, SLC34A2, SLC7A7, SMPD1, SNAPIN, STAT3, STAT5B, TERT, TINF2*, and *ZCCHC8*) was performed on the exome data. Regions of interest across these genes had a minimum coverage of 30x. All detected variants within these genes, including variants of uncertain significance (VUS), were manually reviewed and interpreted for potential clinical relevance.

**Sanger sequencing*:*** Patient consent was obtained before collecting 2 mL of venous blood. DNA samples were isolated using an automated DNA isolation system (EZ2 Connect, Qiagen). Exon 34 of the *LRRK2* gene (NM_198578.4) was amplified using the forward primer 5’-CTA GGC CAC ATG GTT GCT AGA-3’ and the reverse primer 5’-CAG TAG GAG GTT TAC ACT AGA AGC A-3’. PCR amplification was performed using a standard protocol with recombinant Taq DNA polymerase (Thermo Scientific), employing µM primer pairs, 1X (NH4)2SO4 buffer, 2 mM MgCl2, and 0.2 mM dNTP mix. A two-step annealing protocol with temperatures ranging from 60°C to 58°C was applied. Following purification, the amplified fragments (485 bp) were sequenced using Sanger sequencing on an ABI 3500 Genetic Analyzer (Applied Biosystems). The resulting electropherograms were aligned to the GRCh37/hg19 reference genome using the UCSC BLAT tool and subsequently analysed.

**Histopathology:**

Hematoxylin and eosin (H&E) stained histologic slides and corresponding paraffin tissue blocks from seven regions of the explant lungs from index patient III-2 as well as H&E-stained histologic slides and immunohistochemical stains for TTF1 and pancytokeratin AE1/AE3 representing the lung biopsy from the affected brother, patient III-1, were received for histopathologic analysis. Additional Trichrome-VVG stained slides and ABCA3 immunohistochemical stained slides were generated by the Cincinnati Children’s Research Foundation (CCRF) Integrated Pathology Research Facility, Cincinnati, OH. The ABCA3 immunhistochemical stain was generated using a Ventana BenchMark ULTRA automated stainer with EDTA retrieval and hand application of the ABCA3 rabbit polyclonal antibody from Seven Hills Bioreagents (Catalog # WRAB-70565) applied at 1:200 dilution. Microscopic images were acquired using a Nikon DS-Ri2 camera.

**Immunofluorescence:**

Lung tissues were fixed in 10% formalin and embedded in paraffin. Sections were melted at 60^o^C for two hours and rehydrated through xylene and alcohol, and finally in PBS. Antigen retrieval was performed in 0.1 M citrate buffer (pH 6.0) by microwaving. Slides were blocked for 2 hours at room temperature using 4% normal donkey serum (Jackson Immuno Research Laboratories) in PBS containing 0.2% Triton X-100, and then incubated with primary antibodies diluted in blocking buffer for approximately 16 hours at 4^o^C. Primary antibodies included ABCA3 (1:100, Seven Hills Bioreagents), KRT8 (1:100; Developmental Studies Hybridoma Bank), KRT17 (1:100, Santa Cruz Biotechnologies), LAMP3 (1:200, Novus Biologicals) mSFTPB (1:100, in house), and pSFTPC (1:500, Seven Hills Biologicals). Appropriate secondary antibodies conjugated to Alexa Fluor 488, Alexa Fluor 568, or Alexa Fluor 647 (Thermo Fisher Scientific, Jackson Immuno Research) were used at a dilution of 1:200 in blocking buffer for 1 hour at room temperature. Nuclei were counterstained with DAPI (1 μg/ml), (D21490, Thermo Fisher Scientific). Sections were mounted using ProLong Gold antifade reagent, (P36930, Thermo Fisher Scientific) mounting medium with Number 1.5 coverslip.

**Confocal microscopy.**

Tissue sections stained by immunofluorescence were imaged on a Nikon AXR inverted confocal microscope in NSPARC/SR Mode using a PLAN APO λD 100X Oil OFN25 DIC N2 objective. Pixel size = 0.03µm/px. Super resolution, Z stack images were obtained sequentially using channel series and were generated using Nikon NIS-Elements software. Deconvolution was performed on Z stack images by Landweber method using 15 iterations in Nikon NIS-Elements software.

**Transmission electron microscopy:**

Snap-frozen unfixed lung explant tissue from patient III-2 with biallelic *LRRK2* loss-of-function (LoF) variants (obtained from the participating institution) and 250 µm frozen section scrolls from control donor lungs (obtained from LifeCenter Organ Donor Network, Cincinnati, OH and processed in CCRF Integrated Pathology Research Facility, Cincinnati, OH) were fixed in pre-chilled fixative containing 2% paraformaldehyde, 2% glutaraldehyde, and 0.1% calcium chloride in 0.1M sodium cacodylate buffer (pH 7.2) at -20°C. Samples were gradually warmed to 4°C and stored in fresh fixative at 4°C until being process for transmission electron microscopy (TEM) as described previously.^5^ Electron micrographs were acquired using a Hitachi H-7800 TEM (Hitachi High Technologies, Hillsboro, OR) and a BIOSPR16 TEM CCD camera (Advanced Microscopy Techniques, Woburn, MA) within the CCRF Integrated Pathology Research Facility.

**Neutrophil isolation and lysis:**

Neutrophils were isolated from fresh peripheral blood by immunonegative magnetic selection using the EasySep Direct Human Neutrophil Isolation Kit (StemCell Technologies) and processed as previously described. ^6,7^ Peripheral blood collected in K₂EDTA tubes was subjected to sequential magnetic separation according to the manufacturer’s protocol to obtain a neutrophil-enriched fraction.

#### Cells were pelleted by centrifugation at 500g for 5 minutes at room temperature and resuspended in RPMI 1640 medium (Thermo Fisher Scientific). Neutrophils were then incubated with either dimethyl sulfoxide (vehicle control) or 200 nM MLi-2 for 30 minutes. Following treatment, cells were pelleted and lysed in ice-cold lysis buffer supplemented with 0.5 mM diisopropyl fluorophosphate (DIFP) and snap-frozen.

#### Lysates were thawed on ice and clarified by centrifugation at 17,000g for 15 minutes at 4°C. Protein concentration was determined using a bicinchoninic acid (BCA) assay (Thermo Fisher Scientific) according to the manufacturer’s instructions.

**Targeted Mass Spectrometry Analysis:**

Cleared neutrophil lysates (100 μg) were processed using a semi-automated workflow on Myra (Bio Molecular Systems) and Bravo (Agilent) liquid-handling platforms. Proteins were reduced, alkylated, and digested using a single-pot, solid-phase–enhanced sample preparation (SP3) protocol with trypsin/Lys-C. Peptides were acidified, and aliquots were reserved for total Rab10 measurements.

For quantification of phosphorylated Rab10 at threonine 73 and LRRK2 (including Ser910 and Ser935 phosphorylation), peptides were resuspended in immunoprecipitation buffer containing heavy-labeled peptide standards (Table S2) and subjected to automated immunoprecipitation using Protein G magnetic beads conjugated with specific antibodies (Table S3). Enriched peptides were eluted with trifluoroacetic acid.

For total Rab10 measurements, peptides were resuspended in LC buffer containing heavy-labeled peptide standards (Table S2) and loaded onto EvoTips (Evosep) according to the manufacturer’s instructions.

Peptides were analysed using an Evosep One liquid chromatography system coupled to an Orbitrap Exploris 480 mass spectrometer (Thermo Fisher Scientific). Parallel reaction monitoring was performed with predefined precursor isolation windows and optimised collision energies.

Raw data were processed using Skyline (version 24.1.0.199). Light-to-heavy peptide ratios were calculated, and absolute peptide concentrations were determined using external calibration curves. All chromatograms were manually reviewed for peak quality and assignment.

Mass spectrometry proteomics data have been deposited to the ProteomeXchange Consortium via the PRIDE repository. The accession number will be made available at a later stage.

**Urine collection and processing:**

Urine samples were collected and processed as previously described. ^8^ Midstream urine samples were stored at 4°C and processed within 30 minutes. Samples were centrifuged at 2500g for 15 minutes at 4°C, and the supernatant was aliquoted, snap-frozen, and stored at -80°C until analysis.

Lipid extraction and targeted lipidomics: Lipid extraction and targeted lipidomic analysis of bis(monoacylglycerol)phosphate (BMP) species were performed as previously described. ^9^ Briefly, lipids were extracted using a chloroform–methanol–based protocol in the presence of internal standards (SPLASH LIPIDOMIX, Avanti Polar Lipids), followed by liquid chromatography–mass spectrometry analysis.

#### Liquid chromatography was performed using an Agilent 1290 Infinity II system with a C18 column. Mass spectrometry was conducted on an Agilent Ultivo triple quadrupole instrument operating in multiple reaction monitoring (MRM) mode. BMP species were quantified using predefined transitions from ammonium adduct precursor ions to monoacylglycerol fragment ions.

#### Data were processed using Agilent Qualitative and Quantitative Analysis software. Lipid species were quantified based on peak areas, with quality control criteria including a signal-to-noise ratio greater than 3 and linearity (R²>0.95) in pooled quality control samples. BMP levels were normalized to phosphatidylcholine (PC 18:1_18:1) following internal standard normalization.

#### **Plasmids, transient transfection in HEK293 cells, and lysis:**

#### Plasmids used in this study (Table S4) were obtained from MRC PPU Reagents and Services (Dundee, UK). HEK293 overexpression assays were performed as previously described. ^10^

HEK293 cells were maintained in Dulbecco’s Modified Eagle Medium supplemented with 10% fetal bovine serum, 2 mM L-glutamine, and penicillin–streptomycin and cultured at 37°C with 5% CO₂. Cells were transfected at 80–90% confluency with 2 μg of FLAG-tagged LRRK2 constructs or empty vector using polyethylenimine.

Cells were lysed 16 - 20 hours after transfection in ice-cold lysis buffer containing protease and phosphatase inhibitors. Lysates were clarified by centrifugation at 17,000g for 15 minutes at 4°C, and protein concentrations were determined using a bicinchoninic acid assay.

#### **Quantitative immunoblot analysis:**

#### Cell lysates were prepared in LDS sample buffer (Thermo Fisher Scientific) containing β-mercaptoethanol and heat-denatured. Equal amounts of protein (10 μg) were resolved on Bis-Tris gradient gels and transferred to nitrocellulose membranes.

Membranes were blocked in 5% milk and incubated overnight at 4°C with primary antibodies (Table S5), followed by incubation with fluorescent secondary antibodies. Signals were detected using a LI-COR Odyssey CLx imaging system and quantified with Image Studio Lite software. GAPDH was used as a loading control.

#### **Statistical analysis:**

#### Data visualisation and statistical analysis were performed using GraphPad Prism (version 10.5.0). Group comparisons were conducted using two-way analysis of variance (ANOVA) followed by Tukey’s post-hoc multiple comparisons test. Significance levels are indicated as follows: (*p<0.05, **p<0.01, ***p<0.001).

**References**

1.	Peljto AL, Blumhagen RZ, Walts AD, et al. Idiopathic Pulmonary Fibrosis Is Associated with Common Genetic Variants and Limited Rare Variants. Am J Respir Crit Care Med 2023;207(9):1194–1202. DOI: 10.1164/rccm.202207-1331OC.

2.	All of Us Research Program Genomics I. Genomic data in the All of Us Research Program. Nature 2024;627(8003):340–346. DOI: 10.1038/s41586-023-06957-x.

3.	Chen S, Francioli LC, Goodrich JK, et al. A genomic mutational constraint map using variation in 76,156 human genomes. Nature 2024;625(7993):92–100. DOI: 10.1038/s41586-023-06045-0.

4.	Richards S, Aziz N, Bale S, et al. Standards and guidelines for the interpretation of sequence variants: a joint consensus recommendation of the American College of Medical Genetics and Genomics and the Association for Molecular Pathology. Genet Med 2015;17(5):405–24. DOI: 10.1038/gim.2015.30.

5.	Hahn DR, Na CL, Weaver TE. Reserve autophagic capacity in alveolar epithelia provides a replicative niche for influenza A virus. Am J Respir Cell Mol Biol 2014;51(3):400–12. DOI: 10.1165/rcmb.2013-0437OC.

6.	Fan Y, Howden AJM, Sarhan AR, et al. Interrogating Parkinson's disease LRRK2 kinase pathway activity by assessing Rab10 phosphorylation in human neutrophils. Biochem J 2018;475(1):23–44. DOI: 10.1042/BCJ20170803.

7.	Fan Y, Nirujogi RS, Garrido A, et al. R1441G but not G2019S mutation enhances LRRK2 mediated Rab10 phosphorylation in human peripheral blood neutrophils. Acta Neuropathol 2021;142(3):475–494. DOI: 10.1007/s00401-021-02325-z.

8.	Gomes S, Garrido A, Tonelli F, et al. Elevated urine BMP phospholipids in LRRK2 and VPS35 mutation carriers with and without Parkinson's disease. *NPJ Parkinsons Dis* 2023;9(1):52. doi: 10.1038/s41531-023-00482-4.

9.	Dong W, Nyame K, Rawat ES, et al. Robust analytical methods for bis(monoacylglycero)phosphate profiling in health and disease. bioRxiv. February 13, 2025. doi:10.1101/2025.02.13.638174.

10.	Pratuseviciute N, Lis P, Weber S, et al. Functional and structural characterization of LRRK2 p.V1447L in Parkinson’s disease. Mov Disord. 2025;40(10):2251-2256. doi:10.1002/mds.30284.
